# Evaluation of the Abbott Architect, Roche Elecsys and Virtus S1 SARS-CoV-2 antibody tests in community-managed COVID-19 cases

**DOI:** 10.1101/2020.10.27.20220509

**Authors:** Sebastian L. Johnston, Paul F McKay, Tatiana Kebadze, Kai Hu, Karnyart Samnuan, Juliya Aniscenko, Aoife Cameron, Neeta Patel, Paul Randell, Robin J Shattock, Michael R Edwards

## Abstract

**Background:** Antibody testing can help define how protective immunity to SARS-CoV-2 is and how long this immunity lasts. Many antibody tests have been evaluated in hospitalised rather than community based COVID-19 cases. Virtus Respiratory Research Ltd (Virtus) has developed its own quantitative IgM and IgG SARS CoV-2 antibody assay. We report its validation and performance characteristics and compare its performance with the Abbott Architect and Roche Elecsys assays in community COVID cases.

**Methods:** We developed a quantitative antibody test to detect IgM and IgG to the SARS-CoV-2 S1 spike protein (the Virtus test) and validated this test in 107 “true positive” sera from 106 community-managed and 1 hospitalised COVID-19 cases and 208 “true negative” serum samples. We validated the Virtus test against a neutralising antibody test. We determined sensitivities of the Abbott test in the 107 true positive samples and the Roche test in a subset of 75 true positive samples.

**Results:** The Virtus quantitative test was positive in 93 of 107 (87%) community cases of COVID-19 and both IgM and IgG levels correlated strongly with neutralising antibody titres (r=0.75 for IgM, r=0.71 for IgG, *P*<0.0001 for both antibodies). The specificity of the Virtus test was 98.6% for low level antibody positives, 99.5% for moderate positives and 100% for high or very high positives. The Abbott test had a sensitivity of 68%. In the 75 sample subset, the Virtus test was positive in 91%, the Roche test in 69%.

**Conclusions:** The Abbott and Roche tests had sensitives of 68% and 69% respectively in this community set of COVID-19 sera, while the Virtus test had sensitivities of 87% and 91% in the same sample sets. The strong positive correlation with virus neutralization suggests a positive Virtus quantitative antibody test is likely predictive of protective against recurrent COVID-19.

**Funding:** The development of the Virtus test and sample testing with all antibody tests was funded by Virtus Respiratory Research Ltd. The research studies providing 111 of the 208 of the “true negative” samples was supported by MRC Grant numbers MR/M025330/1 and G1100238 and by the National Institute of Health Research (NIHR) Imperial Biomedical Research Centre (BRC), SLJ is a NIHR Emeritus Senior Investigator and is funded in part by European Research Council Advanced Grant 788575 and the Asthma UK Clinical Chair (grant CH11SJ). The views expressed are those of the author(s) and not necessarily those of the NIHR or the Department of Health and Social Care.

## Introduction

The global impact of COVID-19 is immense and the public health threat it poses is unprecedented, immediate and ongoing(1). Public health responses to COVID-19, the disease caused by SARS-CoV-2 infection, depend upon understanding the degree to which SARS-CoV-2 infection results in immune responses that protect people from future infections or illness, and the duration of such protection(2). Individuals who have been infected with SARS-CoV-2 also want to know to what degree and for how long they may be protected. The presence or absence of protective immunity to infection can be assessed by studying antibody levels.

Experience with seasonal coronavirus infections indicates that of higher titres of virus-specific antibody is protective against disease expression upon re-infection. An experimental study with coronavirus 229E demonstrated that 10 of 15 challenged volunteers became infected, and that those who did not become infected had significantly higher serum IgG antibody levels than those that did, indicating that higher levels of antibody were a marker of protection from infection(3). Nine of the 10 infected subjects were re-challenged with the same dose of the same virus approximately one year later, when their antibody levels had declined to around one third of the post-infection peak, but were still higher than their levels before the first virus challenge. Six of the 9 re-challenged subjects became infected (shed virus), but the duration of virus shedding was much shorter than on the first infection (2 vs 6 days) and none of the 9 developed any symptoms (8 of the 10 infected subjects had cold symptoms on the first infection(3). Thus virus shedding and illness were both reduced on re-infection when antibody levels were higher, consistent with antibody being a marker of protection from disease upon re-infection(3), and emphasising the merit of quantitative antibody testing, so that the strength of the immune response can be assessed.

The Abbott Architect SARS-CoV-2 antibody test is a qualitative test that gives a positive/negative result for IgG against the SARS-CoV-2 nucleoprotein. Based on validation in sera taken ≥14 days from symptom onset from 31 cases (severity unspecified) of COVID-19, Abbott reported a sensitivity of 100% (95% confidence interval [CI] 95.89-100.00) and on validation in 997 sera taken pre-COVID-19, a specificity of 99.6% (95% CI: 98.98-99.89)(4). The Roche Elecsys test is a qualitative test that gives a positive/negative result for total antibodies (subtype unspecified) against the SARS-CoV-2 nucleoprotein. Based on validation in sera taken ≥14 days from symptom onset from 29 cases (severity unspecified) of COVID-19, Roche also reported a sensitivity of 100% (95% CI 88.1-100) and on validation in 5,272 sera taken pre-COVID-19, a specificity of 99.81% (95% CI: 99.65-99.91)(5). Public Health England (PHE) evaluated the Abbott and Roche assays in 96 and 93 sera from recovered COVID-19 cases respectively and reported sensitivities of 92.7% (95%CI 85.6-97.0)(6), and 83.9% (95%CI 74.8-90.7)(7), respectively. Both evaluations included hospitalised COVID-19 cases, but precise numbers of hospitalised vs non-hospitalised cases and severities of disease were not reported. Based on these company-reported sensitivities and PHE evaluations, these assays are now very widely used in diagnostic laboratories.

Further reported sensitivities for the Abbott test are 99.1% in hospitalised patients(8), 97.9% in a mix of hospitalised patients and infected healthcare workers(9), 100% in patients, the vast majority of whom were hospitalised(10), 100 % in sera from patients whose disease severity was not specified(11) and 84.2% in patients admitted to a Singapore hospital, all of whom had respiratory symptoms and/or fever, but disease severity was not further reported(12). Reported sensitivities for the Roche test are 98.3% in sera from mostly hospitalised patients(13). Abbott and Roche test sensitivities were 95.4% and 87.9% respectively in sera from a mix of 40 hospitalised (25 in ICU) patients and 28 infected healthcare workers (14), and 94.0% and 97.4% respectively in sera from hospitalised patients in Taiwan(15) and 92.7% and 97.2% in a mix of infected healthcare workers, blood donors and hospitalised patients(16). There is a paucity of data on the sensitivities of these tests in community-managed COVID-19 cases.

Virtus Respiratory Research Ltd (Virtus) has developed its own in-house antibody test quantitating (through use of a standard curve of proprietary pooled positive sera) levels of both IgM and IgG antibodies to the SARS-CoV-2 S1 subunit of the spike protein. We chose the spike protein because it is the major target of neutralising antibodies(17). Virtus has validated this test in sera from 106 community-managed and one hospitalised COVID-19 cases, including asymptomatic cases, as well as against a neutralisation assay. The Abbott and Roche tests were also evaluated in the same panel of sera.

## Methods

### Sample collection for validation of the Virtus SARS-CoV-2 antibody test

Social media approaches were used to invite 107 community cases of COVID-19 to donate blood to Virtus for commercial development of novel COVID-19 antibody tests. Each donor gave signed informed consent for donation of blood for service evaluation of new SARS-CoV-2 antibody testing services and for use/storage of their data. A brief medical history was taken to ascertain type of testing and date of testing used to verify SARS-CoV-2 infection, along with date of illness onset and a brief description of the symptoms manifest and their severity during the COVID-19 illness.

Negative serum samples were 208 samples comprising 111 samples from healthy subjects collected during two research studies conducted at Imperial College London with all 111 samples collected at least 6 months prior to 31^st^ December 2019 and stored long term at −80°C. Informed signed consent for both research studies was obtained, including permission for future transfer of these samples to commercial entities. The samples were transferred to Virtus under a Material Transfer Agreement with Imperial College London. Research Ethics Committee approval reference numbers for these research studies were 15/LO/1666 and 12/LO/1278. Eighty-two true negative serum samples came from healthy subjects taking part in two Virtus commercially funded research studies with all samples collected at least 6 months prior to 31^st^ December 2019 and stored long term at −80°C. Informed signed consent for both research studies was obtained, including permission for future testing of these samples. Ethics approval numbers for these studies were 15/ES/0112 and 18/EM/0311. The final 15 negative serum samples were sera from clinical test samples submitted to Virtus for commercial testing, having previously been determined negative by the Abbott Architect SARS-CoV-2 IgG antibody assay in The Doctors Laboratory commercial testing laboratory.

### The Virtus quantitative antibody test to detect IgM and IgG to the SARS-CoV-2 S1 spike protein

This assay is an enzyme linked immunosorbent assay (ELISA) testing for serum antibody immunoreactivity to the SARS CoV-2 Spike S1 subunit protein. This assay was built in house using commercially sourced mammalian cell expressed S1 spike protein antigen. Antibodies used were goat anti-human IgM and IgG antibodies used at a dilution of 1:1000. Raw data for antibody binding to S1 antigen and no antigen (background) is produced for each assay. Raw data is recorded as an Absorbance (A) value at 450nM. From this, a mean of duplicates is calculated, and the mean value of the blank plate duplicates is subtracted from that of antigen coated plates. The final value is the A450nM value of antibody binding to S1 antigen – no antigen A450nM value. Negative values are converted to a value of 0.000.

The standard curve used a pooled proprietary positive control serum (3 separate Virtus test-positive serum samples pooled into a single sample) serially diluted 1/2-fold over 6 dilutions and run 21 times for the top point and 8 times for the full standard curve to generate mean values for each point on the standard curve to calibrate unknown samples against. For both IgM and IgG, each experimental point (x) on each standard curve is assigned a known value (y), and values of unknown samples are thus determined based on the performance of the standard curve. Results are expressed as A450nM arbitrary units (AU). Intra-assay reproducibility is <5% Coefficient of Variation (CV) in n=188 duplicates, the inter-assay reproducibility is <15% CV in n=76 replicates.

### The SARS-CoV-2 neutralising antibody test

Further validation was performed using a SARS-CoV-2 pseudo-virus neutralisation assay as described(18). This assay utilises a plasmid encoding the SARS-CoV-2 spike protein, along with a lentivirus plasmid encoding other structural proteins required for the lentivirus capsid, and a plasmid encoding a luciferase reporter gene. When introduced by transfection into competent cells, virus-like particles are produced in the supernatant with the lentivirus capsid expressing the SARS-CoV-2 Spike protein. The virus-like particles are functional and infect the Caco-2 cell line used, which expresses the SARS-CoV-2 receptor ACE2(19). Infection is measured by the production of luciferase, indicting virus entry and production of luciferase protein by the luciferase plasmid. When mixed with serum as a dilution series, infection is potentially blocked by serum antibodies, indicating the presence of SARS-CoV-2 Spike protein neutralising antibody. Neutralisation assay data is expressed as an IC_50_ of serum antibody titre. All neutralisation assays were performed by Imperial College London in Professor Robin Shattock’s laboratory.

### Statistical analysis

Data were presented as estimates with 95% CIs. Comparisons between positive and negative samples was performed using the Mann-Whitney-u test. Correlation analyses were performed with Spearman’s correlation. Statistical analyses were performed using Prism 7.04 (GraphPad Software).

## Results

### Validation of the Virtus quantitative antibody test for IgM and IgG antibodies to the SARS-CoV-2 S1 subunit spike protein

#### Clinical description of the Virtus community COVID-19 positive case serum bank

One hundred and seven (107) serum samples were collected between 11^th^ and 28^th^ April 2020 from community-based individuals with previous test-confirmed COVID-19 infection at least 10 days after the earlier of symptom onset or testing positive for SARS-CoV-2. The mean time of blood sampling from symptom onset/test positive was 28 days (range 10-75). Of these 107 confirmed positive samples, 94 were confirmed positive by PCR reported SARS-CoV-2 positive by NHS or commercial testing laboratories, 13 were positive by a variety of commercially sourced lateral flow antibody tests performed on finger prick capillary blood samples and 7 of the 94 were positive by both PCR and lateral flow antibody tests.

Clinical illness severity varied from asymptomatic cases (n=3) through mild/moderate community-managed disease, only a single case was severe enough to require admission to hospital for 5 days, with oxygen requirement, but no intensive care or assisted ventilation.

#### Determination of cut-off values for positivity

After validation using 107 true positive sera and 208 true negative sera, cut-off values in A450nm were chosen that defined the positive and negative population. These cut-off values were chosen to optimise sensitivity and to meet the specificity requirement announced by the Medicines and Healthcare Regulatory Authority (MHRA) that all COVID-19 antibody tests to be sold for testing of the general public should have a minimum specificity of 98%. The cut off value chosen for positivity in the IgM assay was 0.146AU - a value 9.8 times the mean and 4 times the standard deviation (SD) of the true negative samples, while that in the IgG assay was 0.201AU - a value 7.1 times the mean and 3.85 times the standard deviation (SD) of the true negative samples. The absorbance values for the IgM assay are depicted in Figure 1 and the data are summarised in Table1, while those for the IgG assay are in Figure 2 and Table 2.

**Table 1:** Summary of result data for analysis of true positive and true negative samples for IgM antibodies to the SARS-CoV-2 S1 spike protein in the Virtus test.

|  | True +ves | True -ves |
| --- | --- | --- |
| Number of values | 107 | 208 |
| Minimum | 0 | 0 |
| 25% Percentile | 0.19 | 0 |
| Median | 0.325 | 0 |
| 75% Percentile | 0.826 | 0.01375 |
| Maximum | 3.067 | 0.357 |
| Range | 3.067 | 0.357 |
| 95% CI of median |  |  |
| Lower confidence limit | 0.26 | 0 |
| Upper confidence limit | 0.577 | 0 |
| Mean | 0.6273 | 0.01494 |
| Std. Deviation | 0.6616 | 0.03638 |
| Std. Error of Mean | 0.06396 | 0.002522 |
| Lower 95% CI of mean | 0.5005 | 0.009965 |
| Upper 95% CI of mean | 0.7541 | 0.01991 |

**Table 2:** Summary of result data for analysis of true positive and true negative samples for IgG antibodies to the SARS-CoV-2 S1 spike protein in the Virtus test.

|  | True +ves | True -ves |
| --- | --- | --- |
| Number of values | 107 | 208 |
| Minimum | 0 | 0 |
| 25% Percentile | 0.12 | 0 |
| Median | 0.497 | 0 |
| 75% Percentile | 1.346 | 0.04675 |
| Maximum | 2.398 | 0.421 |
| Range | 2.398 | 0.421 |
| 95% CI of median |  |  |
| Lower confidence limit | 0.296 | 0 |
| Upper confidence limit | 0.827 | 0.006 |
| Mean | 0.7464 | 0.02846 |
| Std. Deviation | 0.7067 | 0.05217 |
| Std. Error of Mean | 0.06832 | 0.003617 |
| Lower 95% CI of mean | 0.6109 | 0.02133 |
| Upper 95% CI of mean | 0.8818 | 0.03559 |

**Figure 1:**
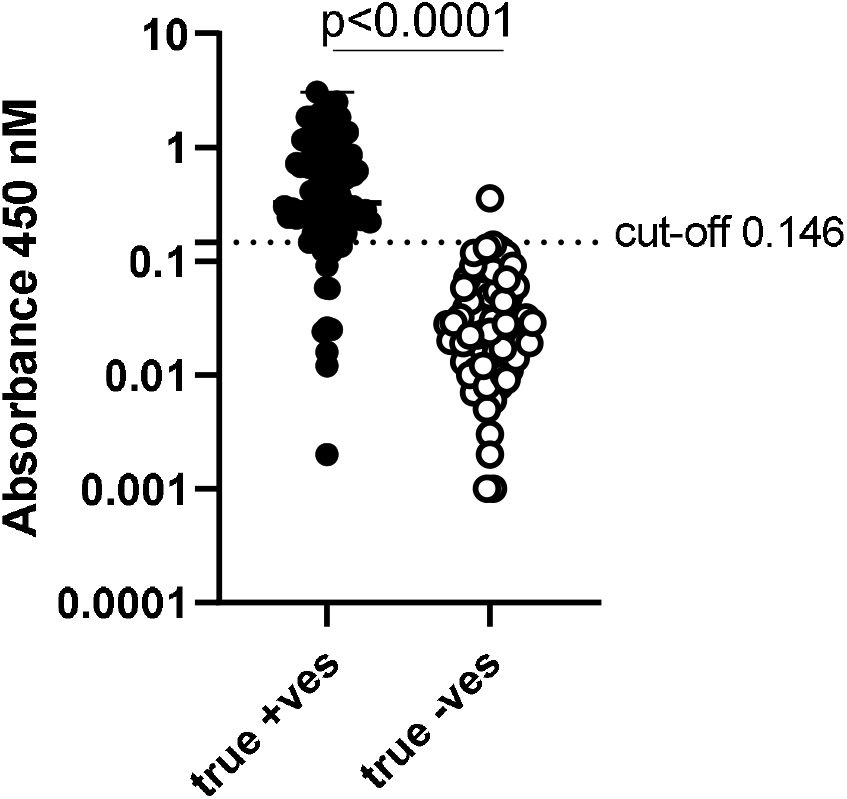
Analysis of true positive and true negative samples for IgM antibodies to the SARS-CoV-2 S1 spike protein in the Virtus test. Absorbance values at 450nM (Log_10_ transformed) results for testing the 107 true positive serum samples (black circles) and 208 true negative serum samples (open circles) for IgM antibody levels. The accepted cut-off for positivity of 0.146 is shown.

**Figure 2:**
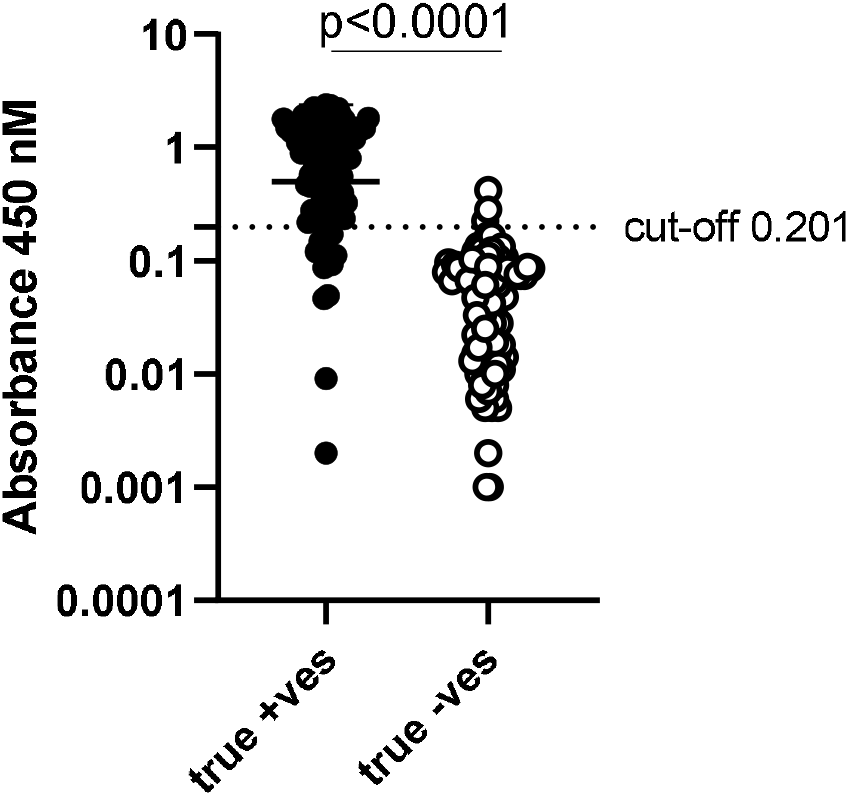
Analysis of true positive and true negative samples for IgG antibodies to the SARS-CoV-2 S1 spike protein in the Virtus test. Absorbance values at 450nM (Log_10_ transformed) results for testing the 107 true positive serum samples (black circles) and 208 true negative serum samples (open circles) for IgM antibody levels. The accepted cut-off for positivity of 0.201 is shown.

#### Determination of cut-off values for low, medium high and very high SARS-CoV-2 antibody positivity

As the Virtus test is a quantitative antibody test giving absorbance values calculated from the standard curve generated from proprietary pooled positive samples, based on A450nM readings, positive samples were further categorised into; low, medium, high or very high positive samples, to permit calculation of specificity for each categorisation. Table 3 describes the cut-off values used to determine the different positive categories.

**Table 3:**
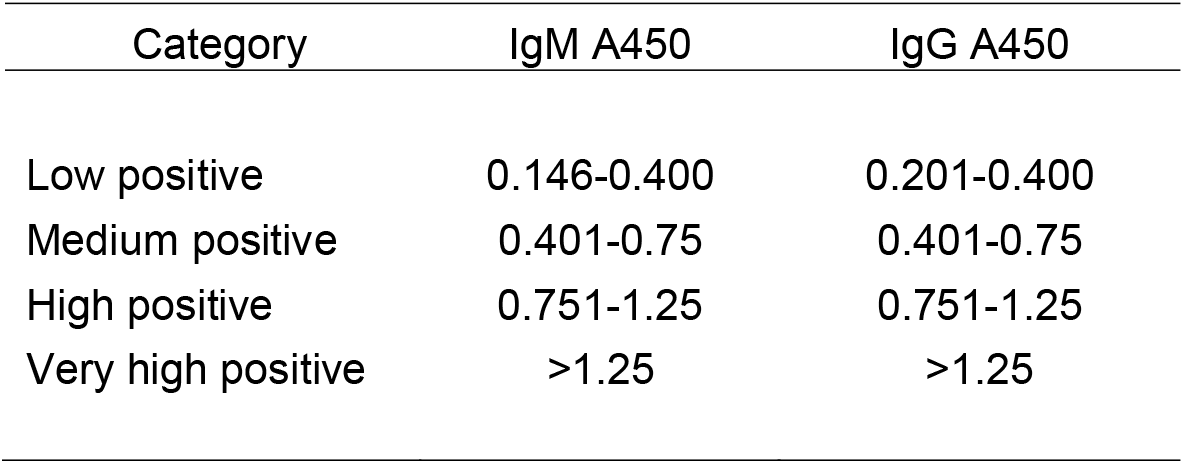
Categorisation of positive samples for IgM and IgG antibodies to the SARS-CoV-2 S1 spike protein in the Virtus test.

| Category | IgM A450 | IgG A450 |
| --- | --- | --- |
| Low positive | 0.146-0.400 | 0.201-0.400 |
| Medium positive | 0.401-0.75 | 0.401-0.75 |
| High positive | 0.751-1.25 | 0.751-1.25 |
| Very high positive | >1.25 | >1.25 |

#### Determination of sensitivity and specificity for low, medium high and very high SARS-CoV-2 antibody positivity

The Virtus quantitative antibody test was positive for IgM and/or IgG in 93 (87%) of the 107 community cases of COVID-19 giving a sensitivity of 86.9% (95% CI 79.0-92.7%).

The specificities for the various cut-offs for low, medium high and very high positivity are given in Table 4.

**Table 4:** Specificities for the categorised positive samples for IgM and IgG antibodies to the SARS-CoV-2 S1 spike protein in the Virtus test.

|  | Low +ve | Medium +ve | High +ve | Very high +ve |
| --- | --- | --- | --- | --- |
| IgM & IgG | 98.6% (3/208) | 99.5% (1/208) | 100% (0/208) | 100% (0/208) |
| 95% CI | 95.8% to 99.7% | 97.4% to 100% | 98.2% to 100% | 98.2% to 100% |

#### Validation of the Virtus SARS-CoV-2 IgM and IgG quantitative serology assay against a virus neutralisation assay

Virus neutralisation tests identify antibodies that are able to block live virus from entering and infecting live cells. To determine whether antibodies detected by the Virtus quantitative antibody test correlate with live virus neutralising activity, we used samples from 32 true positive cases, exhibiting a wide range of A450 values in the Virtus test and compared to data from a pseudo-virus neutralisation assay in which live virus-like particles use the SARS-CoV-2 Spike protein to enter and infect cells(18).

Both IgM and IgG levels detected by the Virtus test correlated strongly with neutralising antibody titres detected in the neutralisation test, with r=0.77, *P*<0.0001 for IgM (Figure 3) and r=0.71, *P*<0.0001 for IgG antibodies (Figure 4).

**Figure 3:**
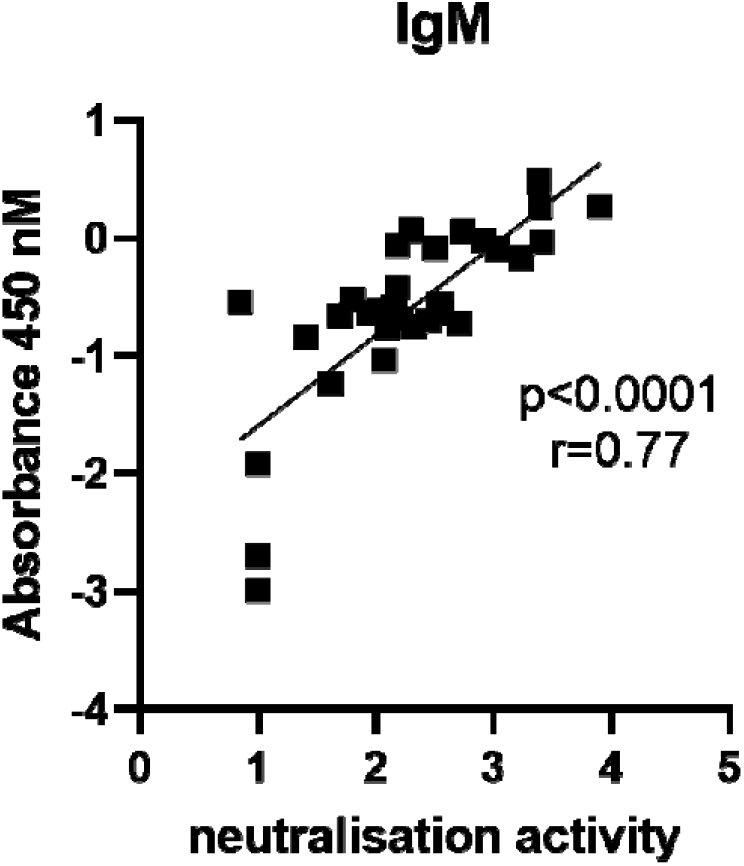
Correlation of true positive samples for IgM antibodies to the SARS-CoV-2 S1 spike protein in the Virtus test with neutralising activity in a neutralising antibody test. Correlation of IgM A450nM results with neutralising activity in the pseudo-virus neutralisation assay. Neutralisation data (x-axis) and SARS-CoV-2 S1 ELISA IgM data (y-axis) are Log_10_ transformed, n=32 samples, r and *P* values are a result of Spearman correlation.

**Figure 4:**
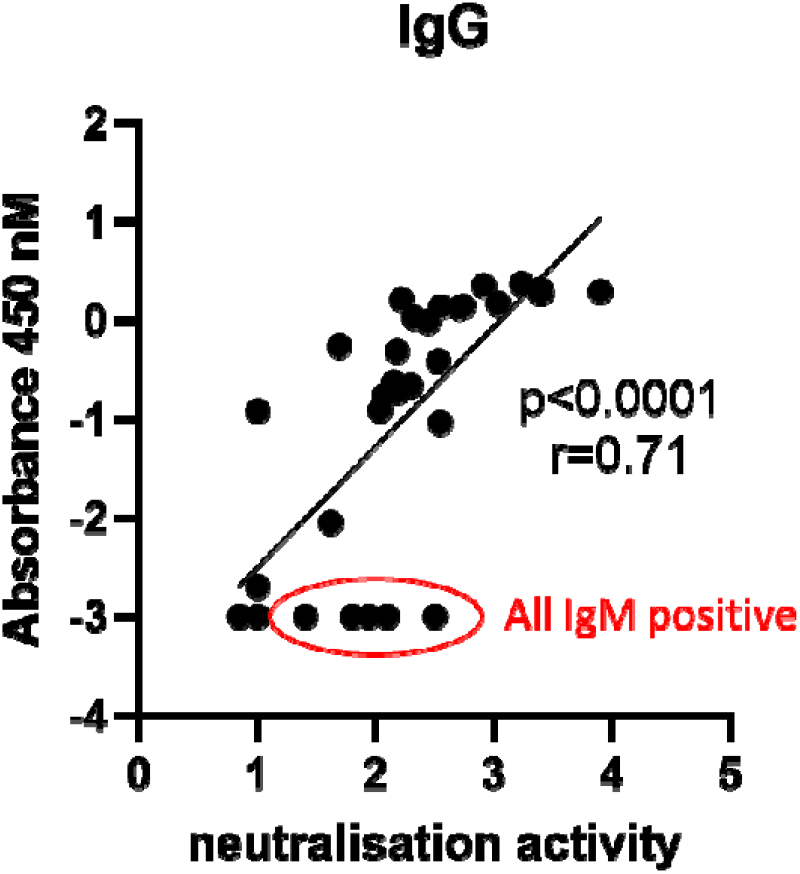
Correlation of true positive samples for IgG antibodies to the SARS-CoV-2 S1 spike protein in the Virtus test with neutralising activity in a neutralising antibody test. Correlation of IgM A450nM results with neutralising activity in the pseudo-virus neutralisation assay. Neutralisation data (x-axis) and SARS-CoV-2 S1 ELISA IgM data (y-axis) are Log_10_ transformed, n=32 samples, r and *P* values are a result of Spearman correlation. Note the 5 samples circled in red with detectable neutralising activity that were negative on IgG testing, all had positive results on testing for IgM.

### Performance evaluation of the Abbott Architect SARS-CoV-2 IgG antibody test in the Virtus community COVID-19 case serum sample bank

All 107 sera from the Virtus COVID-19 test positive community-managed sample set, in which the Virtus test was positive in 86.9% of samples, were analysed in an independent laboratory (North West London Pathology [NWLP] at Charing Cross Hospital using the Abbot Architect test according to the manufacturer’s instructions. The Abbott test was positive in only 73 (68.2%) of these 107 samples, with 20 (18.7%) samples detected by the Virtus test, that were negative on the Abbott test, of these 20 samples, 8 were IgM alone positive samples while 3 were IgG alone and 9 were dual IgM and IgG positive on the Virtus test. There were no samples that were Abbott test positive and Virtus test negative (Supplementary Table 1).

### Performance evaluation of the Roche Elecsys SARS-CoV-2 total antibody test in the Virtus community COVID-19 case serum sample bank

Seventy five of the 107 samples in the Virtus community COVID-19 case serum sample bank underwent testing by NWLP using the Roche assay. The Virtus test was positive in 68 (90.7%) of these 75 samples while the Roche test was positive in only 52 (69.3%) of these 75 samples, with 16 (21.3%) samples detected by the Virtus test, that were negative on the Roche test. As the Roche test is a total antibody test that detects both IgM and IgG, we did not analyse the IgM/IgG positivity of these 16 Roche test negative samples that were positive in the Virtus test. There were no samples that were Roche test positive and Virtus test negative (Supplementary Table 2).

## Discussion

The sensitivity of the Virtus quantitative test for IgM and IgG antibodies to the SARS-CoV-2 spike protein S1 subunit in this set of sera from 106 community managed COVID-19 cases and only a single hospitalised case, was 87%. The Virtus test failed to detect antibody positivity in only 13% of these cases. Despite company claims of 100% sensitivity for both the Abbott and the Roche tests(4, 5), both tests were found in this performance evaluation in community-managed cases of COVID-19 to have substantially lower sensitivities of 68% and 69% respectively. Both Abbott and Roche tests failed to detect antibody positivity in more than 30% of serum samples from community patients previously tested positive for SARS-CoV-2 infection. The majority of these false negative tests in the Abbott and Roche tests were detected as antibody positive in the Virtus test (20/34 samples [59%] for Abbott test false negatives and 16/23 samples [70%] for Roche test false negatives), and all samples which were negative in the Virtus test, were also negative in the Abbott and Roche tests.

Most publications reporting sensitivities for the Abbott Architect and Roche Elecsys SARS-CoV-2 antibody tests have investigated mostly or exclusively hospitalised patients, which in most countries means only people with severe COVID-19, who therefore have high virus loads stimulating very robust immune responses, not unexpectedly resulting in reported sensitivities of 99.1%(8), 97.9%(9), 100%(10, 11), 98.3%(13), 95.4% and 87.9% respectively(14), 94.0% and 97.4% respectively(15) and 92.7% and 97.2% respectively(16). The lowest sensitivity reported is 84.2% in patients admitted to a Singapore Hospital, all of whom had respiratory symptoms and/or fever, but disease severity was not further reported, but possibly was milder in this low prevalence country(12).

There is little data on the sensitivities of antibody tests in community COVID-19 cases, however a pre-print reported sensitivities of both tests in 26 individuals who had SARS-CoV-2 neutralising antibodies detected by in-house neutralisation testing and who were social and working contacts of an index case in a community outbreak in Germany. Only 5 of these 26 individuals were asymptomatic, with the rest having typical symptoms including headache, sore throat, myalgia, cough, fatigue, anosmia, ageusia and other typical symptoms. Twenty 23 (88.5%) of the 26 were also positive for SARS-CoV-2 IgG by immunofluorescence testing. The sensitivity of the Abbott test in these 26 neutralising antibody-positive sera was only 61.5% and that of the Roche test only 65.4%(20). These figures in very similar to the sensitivities in our own community COVID-19 serum panel of 68% for the Abbott test and 69% for the Roche test. Other commonly used tests were also assessed in these 26 neutralising antibody-positive sera, with the Diasorin IgG to S1/S2 test performing similarly to the Abbott at 61.5% positive, while the EUROIMMUN S1 IgA, S1 IgG and combined S1 IgA and/or IgG having positivities of only 46.2%, 46.2% and 53.8% respectively(20).

A low sensitivity for the Abbott test was suggested by personal communications from large numbers of London GPs who had patients whom they were 100% certain had had significant clinical COVID-19 (at a time when no community PCR testing was available in the UK), but who had subsequently tested negative on the Abbott test. Low sensitivity was also strongly suggested by results from the Spanish national serologic survey, in which the Abbott Architect test was used to screen 5118 people who had typical COVID-19 symptoms (anosmia or ageusia, or at least three symptoms among: fever; chills; severe tiredness; sore throat; cough; shortness of breath; headache; or nausea, vomiting, or diarrhoea) at least 14 days prior to sampling, only 18·0% of whom were positive on the Abbott test(21). The authors’ interpretation of the fact that only 18% of symptomatic participants had antibodies against SARS-CoV-2 was that “a sizable proportion of suspected cases might have had symptoms not caused by this coronavirus”(21). We find the suggestion that up to 82% of people, with typical COVID-19 symptoms at a time when SARS-CoV-2 was rampant in Spain, did not have COVID-19, rather implausible. A far likelier explanation in our view is that the Abbott test used had low sensitivity. This interpretation is supported by our finding of 68% sensitivity for the Abbott test in our community cases of COVID-19.

Our findings are also supported by a recent study reporting that 58% of 511 clinical samples that had tested negative on the Abbott test, tested positive on an in-house double binding antigen ELISA (the Imperial Hybrid DABA test), which detects total antibodies to SARS-CoV-2 spike protein receptor binding domain (a subunit of the S1 protein used in the Virtus S1 test)(17).

Our findings have important implications for sero-epidemiologic surveys attempting to define prevalence of past SARS-CoV-2 infections, in which the Abbott and Roche tests have been used to determine prevalence of SARS-CoV-2 antibody positivity. The Abbott test was used in the Spanish National survey(21) and our findings indicate that the prevalences reported therein have been underestimated by at least 30%.

The Virtus test, as well as being the only quantitative test among the three tests evaluated, and thus providing a measure of the strength of immune response generated against SARS-CoV-2, is also the only one of the three tests which was validated against a pseudo-virus neutralisation assay. This assay detects antibodies that are able to block SARS-CoV-2 spike protein-mediated viral entry, and thus antibodies that are functional in preventing live virus infection(18). Given the highly significant and strong correlations observed between IgM and IgG antibodies measured by the Virtus test, and neutralising antibody titres in the same samples, it is highly probable that the antibodies measured by the Virtus test are also likely to be functional, and capable of blocking SARS-CoV-2 spike protein-mediated virus entry. As reported by others(17), this is less likely to be the case for tests like the Abbott and Roche tests which use the nucleoprotein, rather than the surface expressed spike protein. The Virtus test had a substantially better sensitivity that either the Abbott test or the Roche test, detecting 87% compared with 68% and 69% respectively, however it still failed to detect antibody in 13% of this evaluation set of community COVID case sera. One possible explanation for this might be that certain people produce antibodies that bind the SARS-CoV-2 nucleoprotein (the target antigen used by both the Abbot and Roche assay) but not antibodies that bind the SARS-CoV-2 Spike S1 protein (the target antigen in the Virtus assay). The fact that there was not a single Abbott positive, Virtus negative sample, nor a single Roche positive, Virtus negative sample suggests that this is unlikely to be the explanation. An alternative explanation may be that the Virtus test, despite being more sensitive than either the Abbott or Roche tests, may still be less sensitive than optimal. The cut-offs for positivity for the Virtus test were required by the MHRA to be set such that positivity in pre-COVID-19 sera was less than 2%, and therefore specificity >98%. This requirement makes the unproven assumption that there is no, or almost no cross-reactivity in antibody responses between antibodies generated by prior seasonal coronavirus exposure, and antibody responses to SARS-CoV-2. Given that T cell responses to SARS-CoV-2 have been reported in large numbers of studies investigating blood samples taken long before SARS-CoV-2 infected humans(22-27), it seems almost certain that there is significant cross-reactive T cell immunity, and therefore highly likely there will be cross-reactivity in antibody responses as well. The Virtus cut-off for positivity will, by definition, as required by the MHRA, not detect such cross-reactive antibody responses in any more than 2% of people not exposed to SARS-CoV-2. Studies with a quantitative antibody test, such as the Virtus test, measuring antibody levels in large numbers of people on a monthly basis to determine initial as well as long term antibody status, accompanied by regular swab testing (preferably twice weekly, as recurrent infections can be very short-lived(3)) to identify asymptomatic as well as symptomatic SARS-CoV-2 re-infections, will be needed to determine a cut off level for that antibody test that can be definitively shown to provide protection against SARS-CoV-2 re-infection (both asymptomatic as well as symptomatic). It is our belief that that cut-off will be lower than the current cut-off mandated by the MHRA. Such studies are urgently needed.

In conclusion, we report the development and validation of the Virtus S1 antibody test, which is quantitative and validated against a neutralising antibody test, and therefore likely detects antibodies that are functional and protective. Further in a set of sera from recovered COVID-19 cases, all but one of which were community-managed the Virtus test had a sensitivity of 87-91%. In contrast, contrary to company claims of 100% sensitivity, and previous studies reporting mostly >95% sensitivity, the Abbott and Roche tests failed to detect antibody responses in >30% of community cases of SARS-CoV-2 infection. Further refinement and development of antibody tests is needed, including studies investigating quantitative levels of antibody that demonstrably provide protection against SARS-CoV-2 re-infections and disease.

## Data Availability

Data are available from the corresponding author on request

## Author contributions

All authors contributed to the writing of the manuscript and have approved the final version for publication. Paul Randell supervised the Abbott and Roche test analyses and acts as guarantor for the Abbott and Roche test data. Paul McKay, Karnyart Samnuan and Kai Hu performed, and Robin Shattock supervised the neutralising antibody test analyses and act as guarantors for the neutralising antibody data. Michael Edwards, Tatiana Kebadze Juliya Aniscenko, Aoife Cameron and Neeta Patel performed the Virtus test laboratory work. Michael Edwards and Sebastian Johnston led the Virtus assay test development and the performance evaluations of each test and act as guarantors for Virtus data.

## Supplementary appendix

**Supplementary Table 1:**
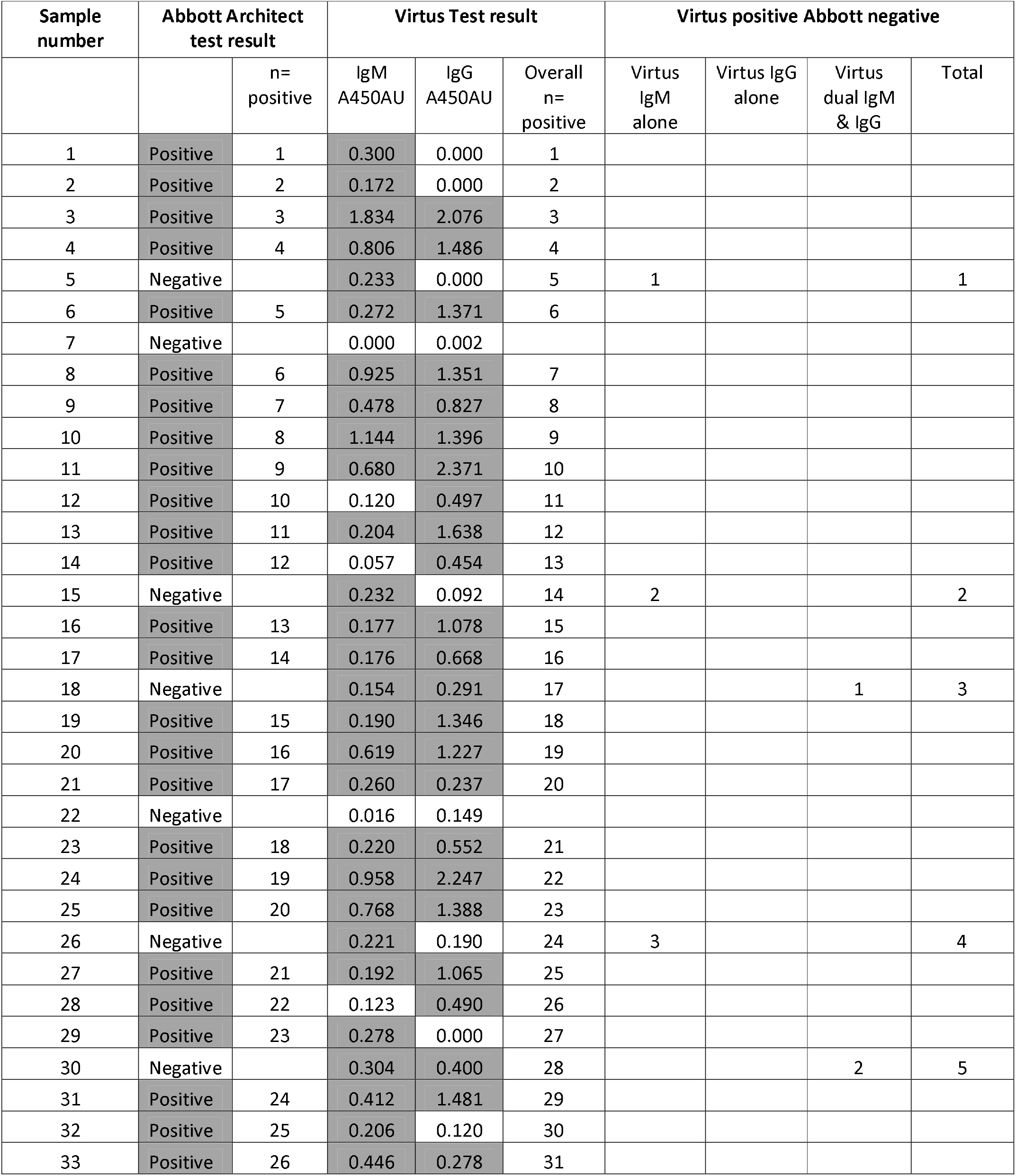
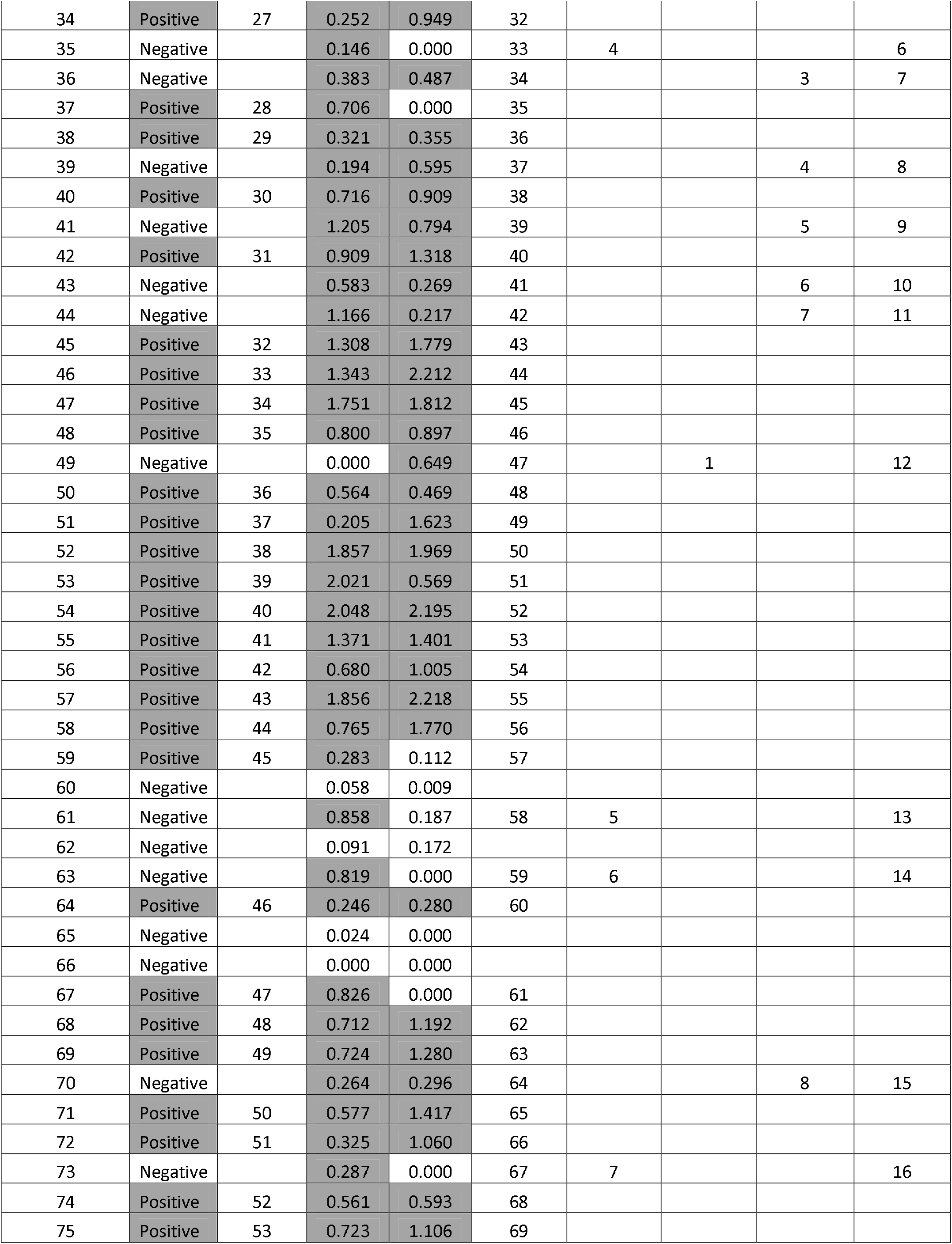
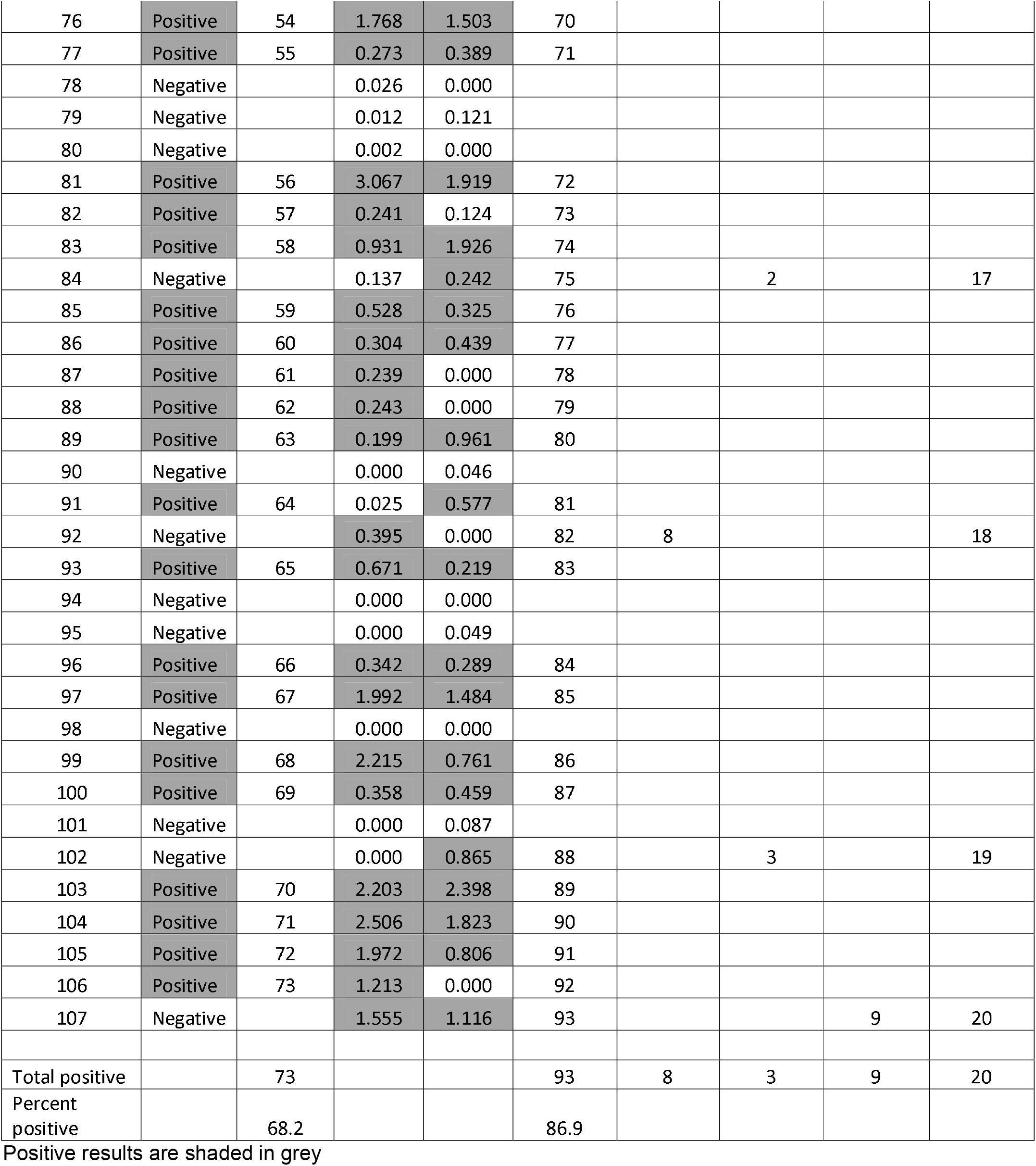
Results of analysis of 107 sera from community based COVID-19 cases for antibodies to SARS-CoV-2 by the qualitative Abbott Architect test and by the Virtus S1 IgM & IgG quantitative test.

**Supplementary Table 2:** Results of analysis of 75 sera from community based COVID-19 cases for antibodies to SARS-CoV-2 by the qualitative Roche Elecsys test and by the Virtus S1 IgM & IgG quantitative test.

| Sample number | Roche Elecsys test result |  | Virtus Test result |  |  |
| --- | --- | --- | --- | --- | --- |
|  |  | n= positive | IgM A450AU | IgG A450AU | Overall n= positive |
| 1 | Negative |  | 0.300 | 0.000 | 1 |
| 4 | Positive | 1 | 0.806 | 1.486 | 2 |
| 5 | Negative |  | 0.233 | 0.000 | 3 |
| 6 | Positive | 2 | 0.272 | 1.371 | 4 |
| 7 | Negative |  | 0.000 | 0.002 |  |
| 8 | Positive | 3 | 0.925 | 1.351 | 5 |
| 9 | Positive | 4 | 0.478 | 0.827 | 6 |
| 10 | Positive | 5 | 1.144 | 1.396 | 7 |
| 11 | Positive | 6 | 0.680 | 2.371 | 8 |
| 12 | Positive | 7 | 0.120 | 0.497 | 9 |
| 13 | Positive | 8 | 0.204 | 1.638 | 10 |
| 14 | Positive | 9 | 0.057 | 0.454 | 11 |
| 15 | Positive | 10 | 0.232 | 0.092 | 12 |
| 16 | Positive | 11 | 0.177 | 1.078 | 13 |
| 17 | Positive | 12 | 0.176 | 0.668 | 14 |
| 18 | Negative |  | 0.154 | 0.291 | 15 |
| 19 | Positive | 13 | 0.190 | 1.346 | 16 |
| 20 | Positive | 14 | 0.619 | 1.227 | 17 |
| 21 | Negative |  | 0.260 | 0.237 | 18 |
| 22 | Negative |  | 0.016 | 0.149 |  |
| 23 | Positive | 15 | 0.220 | 0.552 | 19 |
| 24 | Positive | 16 | 0.958 | 2.247 | 20 |
| 25 | Positive | 17 | 0.768 | 1.388 | 21 |
| 26 | Negative |  | 0.221 | 0.190 | 22 |
| 27 | Positive | 18 | 0.192 | 1.065 | 23 |
| 28 | Positive | 19 | 0.123 | 0.490 | 24 |
| 29 | Positive | 20 | 0.278 | 0.000 | 25 |
| 30 | Negative |  | 0.304 | 0.400 | 26 |
| 31 | Positive | 21 | 0.412 | 1.481 | 27 |
| 32 | Positive | 22 | 0.206 | 0.120 | 28 |
| 33 | Positive | 23 | 0.446 | 0.278 | 29 |
| 34 | Negative |  | 0.252 | 0.949 | 30 |
| 35 | Negative |  | 0.146 | 0.000 | 31 |
| 36 | Positive | 24 | 0.383 | 0.487 | 32 |
| 37 | Positive | 25 | 0.706 | 0.000 | 33 |
| 38 | Positive | 26 | 0.321 | 0.355 | 34 |
| 39 | Negative |  | 0.194 | 0.595 | 35 |
| 40 | Positive | 27 | 0.716 | 0.909 | 36 |
| 41 | Negative |  | 1.205 | 0.794 | 37 |
| 42 | Positive | 28 | 0.909 | 1.318 | 38 |
| 43 | Negative |  | 0.583 | 0.269 | 39 |
| 44 | Negative |  | 1.166 | 0.217 | 40 |
| 45 | Positive | 29 | 1.308 | 1.779 | 41 |
| 46 | Positive | 30 | 1.343 | 2.212 | 42 |
| 47 | Positive | 31 | 1.751 | 1.812 | 43 |
| 48 | Positive | 32 | 0.800 | 0.897 | 44 |
| 49 | Positive | 33 | 0.000 | 0.649 | 45 |
| 50 | Positive | 34 | 0.564 | 0.469 | 46 |
| 51 | Positive | 35 | 0.205 | 1.623 | 47 |
| 52 | Positive | 36 | 1.857 | 1.969 | 48 |
| 53 | Positive | 37 | 2.021 | 0.569 | 49 |
| 54 | Positive | 38 | 2.048 | 2.195 | 50 |
| 55 | Positive | 39 | 1.371 | 1.401 | 51 |
| 56 | Positive | 40 | 0.680 | 1.005 | 52 |
| 57 | Positive | 41 | 1.856 | 2.218 | 53 |
| 58 | Positive | 42 | 0.765 | 1.770 | 54 |
| 59 | Positive | 43 | 0.283 | 0.112 | 55 |
| 60 | Negative |  | 0.058 | 0.009 |  |
| 61 | Negative |  | 0.858 | 0.187 | 56 |
| 62 | Negative |  | 0.091 | 0.172 |  |
| 63 | Negative |  | 0.819 | 0.000 | 57 |
| 64 | Positive | 44 | 0.246 | 0.280 | 58 |
| 65 | Negative |  | 0.024 | 0.000 |  |
| 66 | Negative |  | 0.000 | 0.000 |  |
| 67 | Positive | 45 | 0.826 | 0.000 | 59 |
| 68 | Positive | 46 | 0.712 | 1.192 | 60 |
| 69 | Positive | 47 | 0.724 | 1.280 | 61 |
| 70 | Positive | 48 | 0.264 | 0.296 | 62 |
| 101 | Negative |  | 0.000 | 0.087 |  |
| 102 | Negative |  | 0.000 | 0.865 | 63 |
| 103 | Positive | 49 | 2.203 | 2.398 | 64 |
| 104 | Positive | 50 | 2.506 | 1.823 | 65 |
| 105 | Positive | 51 | 1.972 | 0.806 | 66 |
| 106 | Negative |  | 1.213 | 0.000 | 67 |
| 107 | Positive | 52 | 1.555 | 1.116 | 68 |
| Total positive |  | 52 |  |  | 68 |
| Percent positive |  | 69.3 |  |  | 90.7 |

Positive results are shaded in grey

